# A Novel Digital Phenotype of Physical Activity: Associations with Heart Rate Fragmentation

**DOI:** 10.64898/2026.10.08.26365083

**Authors:** Byung Joon Pae, Ben Cousineau, Zhaohua Zeng, Yuchen Li, Lejy Mathew, Sarah Chang, Katalina De Leon, Laila Wilson, Abigail Alexander, Eric Santana, Nikila Alagar, Kay Colon-Motas, Abdulkareem Murtala, Sajjad Karimi, Qiao Li, Reza Sameni, Yi-An Ko, Felipe Lobelo, Rachel Lampert, J. Douglas Bremner, Viola Vaccarino, Gari D. Clifford, Amit J. Shah

**Affiliations:** Department of Epidemiology, Rollins School of Public Health, Emory University, Atlanta, GA, USA; Department of Medicine, School of Medicine, Emory University, Atlanta, GA, USA; Morehouse School of Medicine, Atlanta, GA, USA; Department of Biomedical Informatics, School of Medicine, Emory University, Atlanta, GA, USA; Wallace H. Coulter Department of Biomedical Engineering, Georgia Institute of Technology and Emory University, Atlanta, GA, USA; Department of Biostatistics and Bioinformatics, Rollins School of Public Health, Emory University, Atlanta, GA, USA; Hubert Department of Global Health, Rollins School of Public Health, Emory University, Atlanta, GA, USA; Section of Cardiovascular Medicine, Department of Internal Medicine, Yale School of Medicine, New Haven, CT, USA; Department of Psychiatry and Behavioral Sciences, School of Medicine, Emory University, Atlanta, GA, USA; Atlanta VA Health Care System, Decatur, GA, USA; Department of Anesthesiology and Critical Care Medicine, School of Medicine, Johns Hopkins University, Baltimore, MD, USA; Department of Biomedical Engineering, Whiting School of Engineering, Johns Hopkins University, Baltimore, MD, USA

## Abstract

**Background:** Heart rate fragmentation (HRF) has emerged as a novel marker of sinoatrial instability and neuroautonomic dysregulation and is a predictor of incident cardiovascular disease (CVD) events and cardiovascular mortality. Leveraging heart rate fragmentation as a novel measure may help us better understand the prognostic significance of physical activity. Physical activity (PA) patterns are multidimensional and difficult to study holistically, however. We sought to examine for novel PA clusters in a cohort of veteran men and evaluate their prognostic significance based on their associations with neuroautonomic dysregulation.

**Methods:** In this cross-sectional study, we examined 602 male veterans from the Vietnam Era Twins Study of Electrophysiology in Posttraumatic Stress Disorder (VETS-EP) who were enrolled between 2022 and 2026. Accelerometry data were used to derive physical activity parameters across 6 domains: volume, sedentary behavior, intensity, diurnal pattern, fragmentation, and variability. Heart rate fragmentation was assessed using percentage of inflection points (PIP), inverse average length of segments (IALS), percentage of short segments (PSS), and percentage of alternating segments (PAS). Gaussian mixture models were used to derive digital phenotypes of physical activity, and associations with heart rate fragmentation were assessed using linear mixed models.

**Results:** Participants had a mean age of 73.8 ± 2.9 years and 93.2% reported White race. Two overlapping phenotypes of physical activity patterns were identified: a compact-early phenotype, characterized by earlier activity timing and slightly greater overall activity, and a diffuse-late phenotype, characterized by later activity timing and greater heterogeneity in activity volume. In fully adjusted models, the diffuse-late phenotype was associated with higher PIP, the primary outcome (β: 1.199, 95% CI: 0.034 to 2.363, p-value: 0.044), and higher IALS (β: 0.01210, 95% CI: 0.00048 to 0.02373, p-value: 0.041), but not PSS or PAS, compared with the compact-early phenotype.

**Conclusions:** In older male veterans, Gaussian mixture model phenotyping identified two overlapping physical activity phenotypes, differing mainly in diurnal timing and in the dispersion of overall activity, that were associated with modest differences in heart rate fragmentation. These phenotypes provide a framework for capturing complex physical activity patterns. Future prospective studies representing diverse populations are needed to validate such phenotypes.

## Introduction

Cardiovascular disease (CVD) remains a leading cause of death globally.^1,2^ Estimates from the Global Burden of Disease study indicate that ischemic heart disease was the leading age-standardized cause of death worldwide in 1990, 2019, and 2021.^1^ Furthermore, projections of CVD from 2025 to 2050 suggest a 90.0% increase in prevalence, 73.4% increase in crude mortality, and 35.6 million anticipated cardiovascular deaths in 2050.^2^ Physical activity is among the modifiable factors most strongly and consistently associated with CVD incidence and mortality,^3, 4^ and physical activity–based interventions are low-cost and scalable worldwide; research characterizing how physical activity patterns relate to CVD risk is therefore critical.

In recent years, heart rate fragmentation has emerged as a novel marker of sinoatrial instability and neuroautonomic dysregulation and a predictor of incident cardiovascular events and cardiovascular mortality.^5, 6^ Unlike heart rate variability, which quantifies the magnitude of beat-to-beat variation,^7^ heart rate fragmentation characterizes the frequency and pattern of directional changes in cardiac inter-beat intervals and may therefore capture the dysregulation of the neuroautonomic-electrophysiologic control system.^5^ In older adults, erratic, non-autonomic fluctuations in sinus rhythm can inflate short-term heart rate variability indices, which may then be misinterpreted as enhanced vagal tone;^5^ heart rate fragmentation quantifies these fluctuations directly and, unlike short-term heart rate variability, increases consistently with age.^5, 6^ Thus, heart rate fragmentation may be appropriate for aging populations.

Autonomic dysfunction, characterized by reduced parasympathetic and heightened sympathetic activity, is a key pathophysiological pathway linking physical inactivity to CVD, and physical activity is well established to improve cardiac autonomic function.^8^ However, age-related deterioration of cardiac rhythm regulation also involves the sinoatrial node itself, which heart rate fragmentation is thought to reflect in addition to dysautonomia,^5^ and heart rate fragmentation predicts adverse cardiovascular events and is well suited to older populations.^5, 6^ Examining physical activity in relation to heart rate fragmentation may therefore clarify whether activity patterns relate to sinoatrial abnormalities in addition to dysautonomia, using an outcome with prognostic relevance for older adults. Moreover, prior studies have focused on traditional volume-based metrics of physical activity,^3, 4^ but there is growing evidence suggesting the potential utility of pattern-based metrics of physical activity.^9, 10, 11, 12^ These alternative metrics evaluate how physical activity is structured and accumulated, encompassing aspects such as fragmentation, diurnal patterns, and temporal distribution of physical activity. To date, however, these investigations have largely evaluated individual pattern-based metrics in isolation and may not account for their combined, multi-dimensional architecture. A multi-dimensional approach that integrates both volume-and pattern-based metrics into a digital phenotype may better capture the complex relationship between physical activity and heart rate fragmentation. Because physical activity patterns are inherently complex, untangling such patterns is needed so that lifestyle modification programs can give patients specific, actionable recommendations, such as when and how activity is accumulated across the day, rather than volume targets alone. Such investigation may help identify specific phenotypes of physical activity patterns that could be promoted for the prevention of CVD and targeted for personalized exercise interventions.

This study aims to develop novel digital phenotypes of physical activity patterns and investigate their associations with four heart rate fragmentation metrics: percentage of inflection points (PIP), inverse average length of segments (IALS), percentage of short segments (PSS), and percentage of alternating segments (PAS). Because PIP and IALS capture the frequency and density of directional changes in cardiac inter-beat intervals, while PSS and PAS capture the accumulation of short and alternating runs of heart rate acceleration and deceleration,^5^ these metrics offer complementary physiological insights. We hypothesize that phenotypes characterized by more favorable physical activity patterns will exhibit lower heart rate fragmentation, particularly lower PIP, which we designated as the primary outcome.

## Methods

### Study Participants

Data were obtained from the Vietnam Era Twins Study of Electrophysiology in Posttraumatic Stress Disorder (VETS-EP), an ongoing longitudinal cohort study aimed at investigating the relationship between posttraumatic stress disorder and risk of sudden cardiac death using electrocardiographic measures. Participants were enrolled between 2022 and 2026 and recruited via phone and mail from the Vietnam Era Twin Registry, a national registry consisting of male twins who served in the U.S. military between 1964 and 1975.^13^ From a total of 943 participants, those with missing physical activity (n=159) were excluded first, then sequentially heart rate fragmentation (n=171), and covariate data (n=11) were excluded. A total of 602 participants were included in the current study’s complete-case analysis (Figure 1). Because heart rate fragmentation processing was not yet complete for the full cohort at the time of this analysis, missing heart rate fragmentation data primarily reflects recordings that had not yet been processed; the present report therefore describes interim results from the ongoing study rather than the final dataset. All participants provided informed consent, and the research protocol was approved by the Institutional Review Board at Emory University.

**Figure 1.**
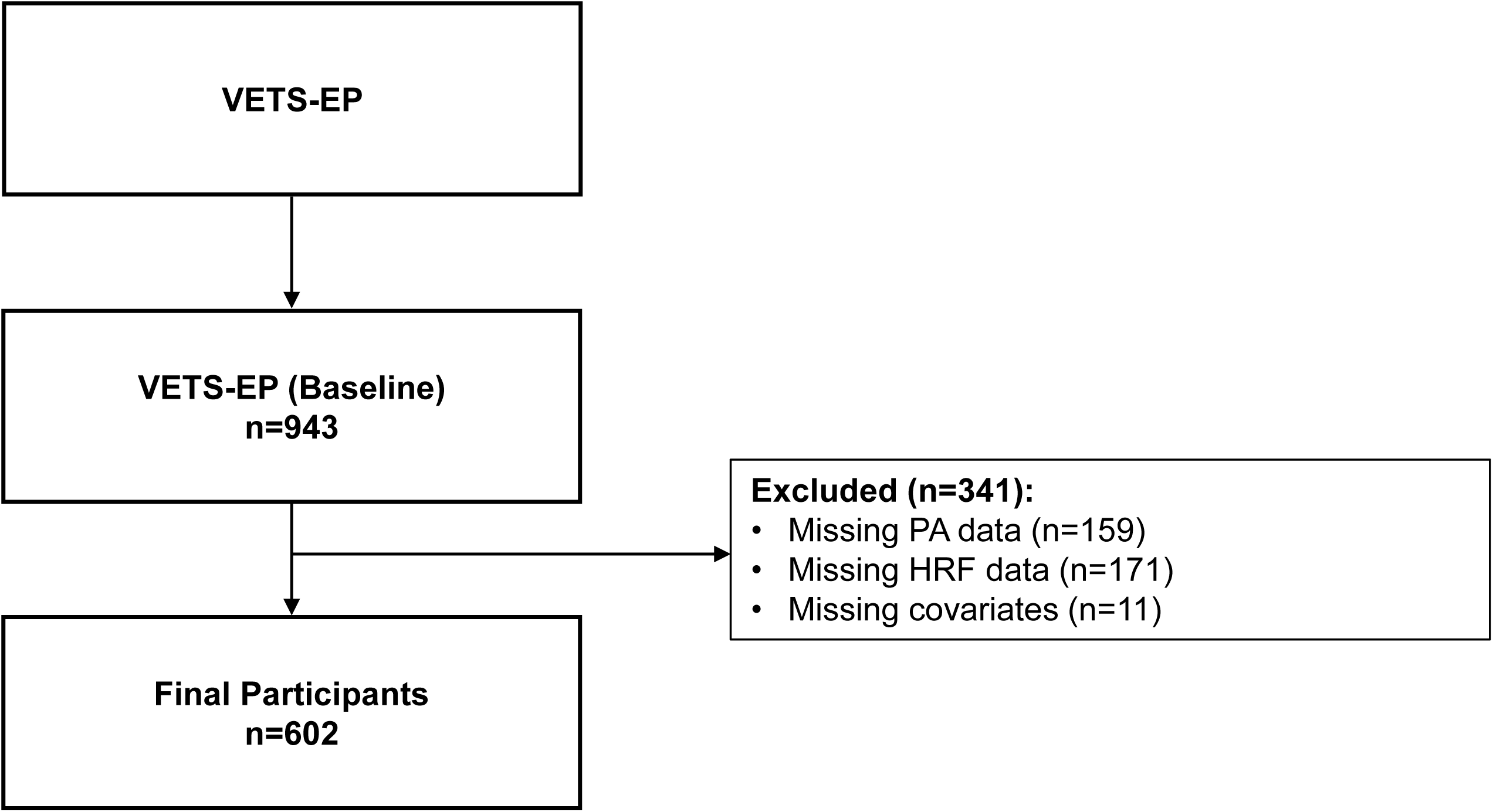
Flowchart of the selection of study participants. VETS-EP, Vietnam Era Twins Study of Electrophysiology in Posttraumatic Stress Disorder; PA, physical activity; HRF, heart rate fragmentation.

### Physical Activity

#### Data Collection

Physical activity was measured using tri-axial, wrist-worn accelerometers (ActiGraph wGT3X-BT; ActiGraph, Pensacola, FL). Acceleration data were sampled at 100 Hz. Participants were instructed to wear the accelerometer continuously for 7 days on their non-dominant wrist, during the same 7-day period as the ambulatory electrocardiogram recording.

#### Data Processing & Feature Extraction

To extract both volume- and pattern-based metrics and ensure compatibility with legacy and current literature, raw accelerometry data were processed using two complementary software pipelines. The ActiLife software (Version 7.3.2; ActiGraph) was used to extract count-based metrics with an epoch length of 10 seconds, and data were scored with the “Worn on Wrist” setting enabled, which applies ActiLife software’s scaling algorithm to convert wrist counts into waist-equivalent counts, with the low-frequency extension filter disabled. The R-based GGIR package (Version 3.3.6)^14^ was used to extract raw acceleration metrics with an epoch length of 5 seconds.

For detecting non-wear time, ActiLife used the Choi (2011) method,^15^ while GGIR evaluated the standard deviation and value range of raw acceleration across rolling 60-minute windows.^16^ Data inclusion thresholds were tailored to the specific output stream. For variables derived from the ActiLife software and GGIR Part 2 output, a valid wear day was defined as having at least 10 hours of wear time, and participants with at least 4 valid wear days were included in the analysis.^17^ For GGIR Part 5 outputs, the default minimum wear time threshold of 16 hours was applied. For GGIR Part 6 outputs, where a daily minimum wear time threshold is not enforced, participants were included if they had at least 4 valid wear days. Only participants with at least 4 valid wear days across all output streams were included in the analysis; otherwise, participants were classified as having missing physical activity data and excluded from the analysis.

The ActiLife software was used to derive daily average step counts, percentage of time spent in sedentary behavior (< 100 counts/minute), and percentage of time spent in moderate-to-vigorous physical activity (MVPA) (≥ 1952 counts/minute). Physical activity intensities were defined using the Freedson Adult (1998) cut points.^18^ The ActiLife software automatically scaled the 60-second intensity cut points to match the 10-second epoch length for intensity classification.

GGIR was used to process raw acceleration data and derive features based on Euclidean Norm Minus One (ENMO) across 3 output streams for acceleration summaries (Part 2), behavioral activity patterns (Part 5), and circadian rhythm metrics (Part 6). Part 2 was used to derive 24-hour full-recording mean ENMO^14, 16^ and daily average intensity gradient (IG) of ENMO.^19^ Part 5 was used to derive daily averages of active-to-sedentary transition probability (ASTP), mean inactive bout length, and number of sedentary bouts lasting at least 30 minutes. The Part 5 algorithm was configured with thresholds of 40 mg for light activity, 100 mg for moderate activity, and 400 mg for vigorous activity.^20^ Inactivity was defined as acceleration less than 40 mg during waking hours.^20^ Because wrist-worn accelerometers cannot assess posture, inactivity may have included sitting, reclining, and standing still. Part 5 outputs were based on the wake-to-wake time window, and all variables derived from Part 5 were calculated exclusively during waking hours and averaged across all valid wear days. Waking and sleeping hours were identified by using the default Heuristic algorithm looking at the Distribution of Change in Z-Angle.^21^ Part 6 was used to derive inter-daily stability (IS), intra-daily variability (IV), and the timing of the most active consecutive 5 hours (M5) and least active consecutive 5 hours (L5). Both M5 and L5 timing represented the number of hours relative to the previous midnight. All variables derived from Part 6 were analyzed across the full time series.

#### Physical Activity Domains

A priori domains of physical activity, including total volume (step count, ENMO),^3, 4, 22^ sedentary behavior (time spent in sedentary behavior, number of sedentary bouts ≥ 30 minutes),^23, 24^ intensity (time spent in MVPA, IG),^9, 23^ diurnal pattern (M5 timing, L5 timing),^10, 25^ fragmentation (ASTP, mean inactive bout length),^12, 26^ and variability (IS, IV)^10, 25^ were considered for developing the multi-dimensional digital phenotypes of physical activity patterns based on established associations with health outcomes. To ensure equal domain representation, each domain included 2 representative variables, yielding a total of 12 physical activity variables.

### Heart Rate Fragmentation

Ambulatory electrocardiograms were recorded for 7 days, with 3 channels and a sampling frequency of 1000 Hz (Bittium Faros 360; Bittium Corporation, Oulu, Finland). To record cardiographic electrical activity under free-living conditions, participants were instructed to wear the device continuously throughout standard daily routines with the exception of water activities. Raw electrocardiogram waveforms were processed using the PhysioNet Cardiovascular Signal Toolbox for QRS detection, artifact correction, implausible interval exclusion, and beat-by-beat annotation.^27, 28^

Segments with a signal quality index (SQI) below 0.7 were excluded, as were segments classified as atrial fibrillation by the toolbox’s atrial fibrillation detection algorithm. Atrial fibrillation-flagged segments were excluded, rather than whole participant exclusion. Normal and ectopic beats were not explicitly differentiated; instead, RR intervals differing by more than 20% from the preceding interval were removed, which excludes most ectopic beats and their compensatory pauses. Paced rhythms were managed via the toolbox’s automated preprocessing routines, utilizing supplied annotation files to flag and exclude non-sinus beats, alongside interval-level artifact and outlier filtering. Heart rate fragmentation was quantified from the resulting RR interval sequences using 4 complementary metrics: PIP, the percentage of intervals at which the sign of successive RR interval differences changes (inflection points); IALS, the inverse of the average length of segments of consecutive accelerations or decelerations; PSS, the percentage of intervals in short segments (fewer than 3 intervals); and PAS, the percentage of intervals in alternation segments of at least 4 intervals in which successive differences alternate in sign.^5^ Because these metrics characterize the frequency and pattern of directional changes in cardiac inter-beat intervals, they may therefore capture the dysregulation of the neuroautonomic-electrophysiologic control system, with higher values reflecting greater fragmentation.^5^ PIP and IALS summarize the overall frequency of directional changes, whereas PSS and PAS characterize specific fragmentation patterns (short runs and sustained alternation, respectively); including all 4 metrics therefore allows assessment of whether associations reflect overall fragmentation or particular fragmentation patterns. Because PIP is the most general of these metrics, and IALS, PSS, and PAS are based on segments delimited by the same inflection points, PIP was designated as the primary outcome, with IALS, PSS, and PAS considered secondary outcomes. All 4 metrics were calculated across the full time series of electrocardiogram monitoring, including periods of both activity and rest.^27, 28^

### Covariates

Covariates included age (years), race (White, Black, and Other), education (Elementary/Middle School, High School, Vocational/Technical School, Some College, College Degree, Post-Graduate College, and Advanced Degree), body mass index (BMI) (kg/m^2^), smoking (Current, Former, or Never), drinking (days of alcohol consumption within last month), hypertension, diabetes, and history of any composite cardiovascular disease (CVD), defined as previous myocardial infarction, revascularization, heart failure, arrhythmia, stroke, or peripheral artery disease. All education categories were retained to reduce residual confounding. Data regarding age, race, education, BMI, smoking, and drinking were self-reported and collected through baseline interviews. Clinical surveys and physical examinations were conducted to collect data for hypertension, diabetes, and CVD. Data collected from clinical surveys were confirmed through medical record reviews of outpatient primary care and cardiology visits. Lifetime depression and posttraumatic stress disorder were also collected from structured clinical interviews and were included for descriptive purposes given their established associations with both physical activity and CVD; however they were not included as covariates in the primary models to avoid potential overadjustment bias, as physical activity may exert a protective causal effect on depression,^29^ positioning these psychiatric conditions as downstream consequences or parallel outcomes rather than upstream confounders.

### Statistical Analysis

#### Unsupervised Machine Learning Phenotyping

The distributions of all extracted physical activity variables were assessed, and log-transformations were performed for two variables with an absolute skewness value greater than 1 (ENMO and inactive bout length). To account for differences in scale and units, z-score transformations were applied to standardize the physical activity variables. Principal component analysis was performed on the standardized physical activity variables, and varimax rotation was applied for improved interpretability. The optimal number of components to retain was determined using Horn’s parallel analysis.^30^ A total of 5000 iterations was used to produce stable estimates, and the empirical eigenvalues were compared against the 95^th^ percentile of the random eigenvalues.^31^

The resulting two principal component scores were used as inputs for the cluster analysis. First, agglomerative hierarchical clustering using Euclidean distance and Ward’s minimum variance method was performed to evaluate the dendrogram and establish candidate cluster values (k).^32^ Next, Gaussian mixture models were fitted across candidate values of k, utilizing agglomerative hierarchical clustering to initialize parameter estimates for the Expectation-Maximization algorithm.^33^ Model selection across candidate k values was determined using the Bayesian Information Criterion (BIC), with the optimal BIC determining the final number of phenotypes. All possible parameterizations were compared. To address potential non-independence of co-twins, we identified 314 independent participants whose co-twins were not present in the dataset and conducted the same unsupervised machine learning phenotyping pipeline. Participants were deterministically assigned to a single phenotype based on maximum posterior probability. Model classification quality was evaluated using assignment probabilities. Because Gaussian mixture models can identify clusters that differ in dispersion as well as location, the non-linearity of the phenotype structure was evaluated by visual inspection of the cluster assignment plot, specifically whether phenotype membership varied monotonically along each principal component. The resulting digital phenotypes of physical activity patterns were characterized and visualized through a cluster assignment plot and standardized z-score heatmap.

#### Associations with Heart Rate Fragmentation

Baseline characteristics for all participants were summarized and compared between the digital phenotypes of physical activity patterns. Continuous variables were compared using Student’s t test or Mann-Whitney U test, depending on normality (for skewed variables, both are presented in Table 1, and the Mann-Whitney U test was considered primary), and categorical variables were compared using the χ2 test. For continuous variables with unequal variances, Satterthwaite’s approximation was utilized. To evaluate component-specific covariate distributions, baseline characteristics were additionally summarized by tertiles of the retained components. Continuous variables were compared using Analysis of Variance (ANOVA) Overall F test or Kruskal-Wallis test, depending on normality, and categorical variables were compared using the χ2 test. Linear mixed models with a random intercept for twin ID were utilized to assess cross-sectional associations between digital phenotypes of physical activity patterns and heart rate fragmentation, with phenotype treated as a categorical variable. Models were sequentially adjusted for a priori selected sets of covariates to evaluate their influence on the associations with fragmentation parameters. Model 1 was a crude model. Model 2 adjusted for age, race, education, BMI, smoking, and drinking. Model 3 additionally adjusted for history of any composite CVD.

**Table 1.**
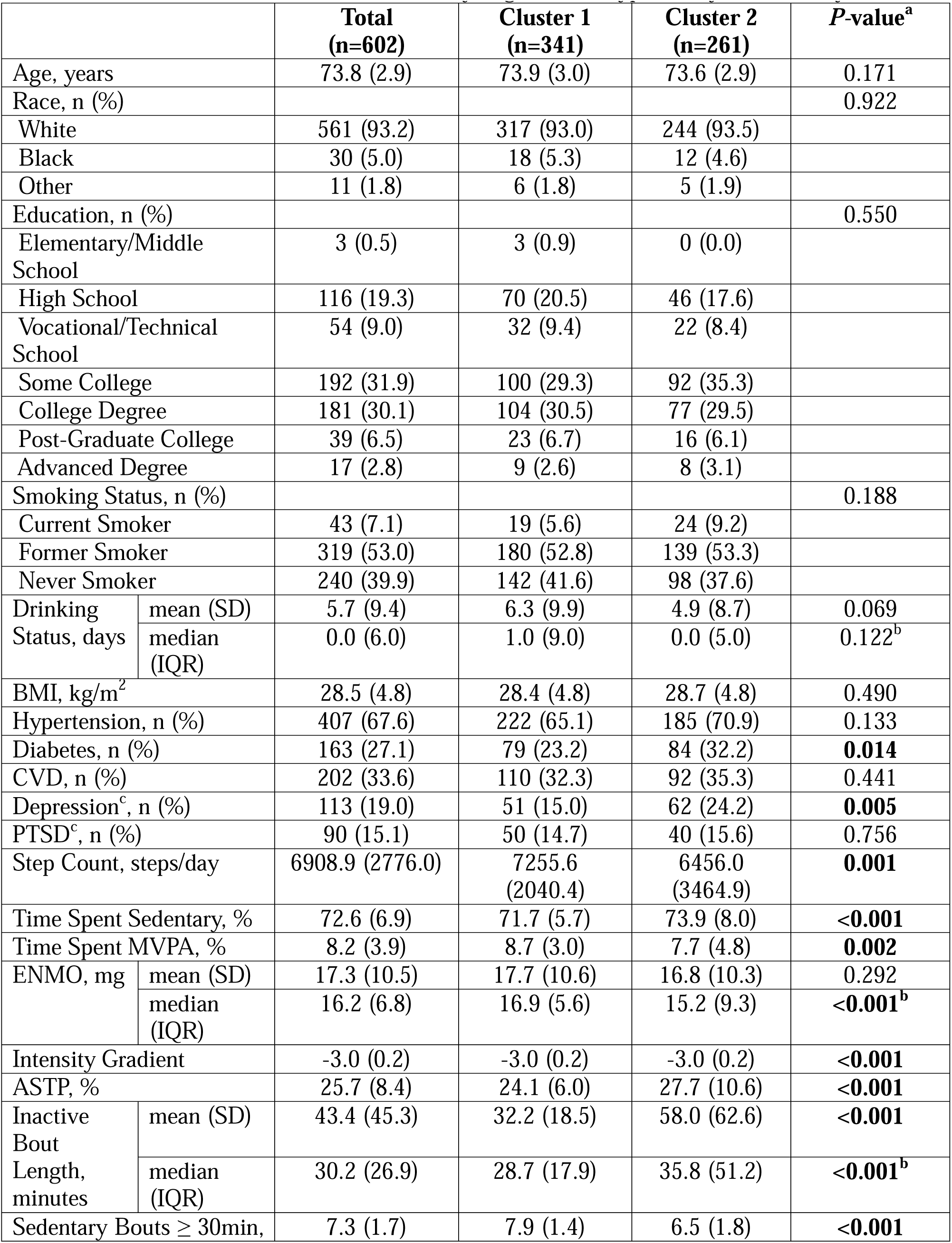
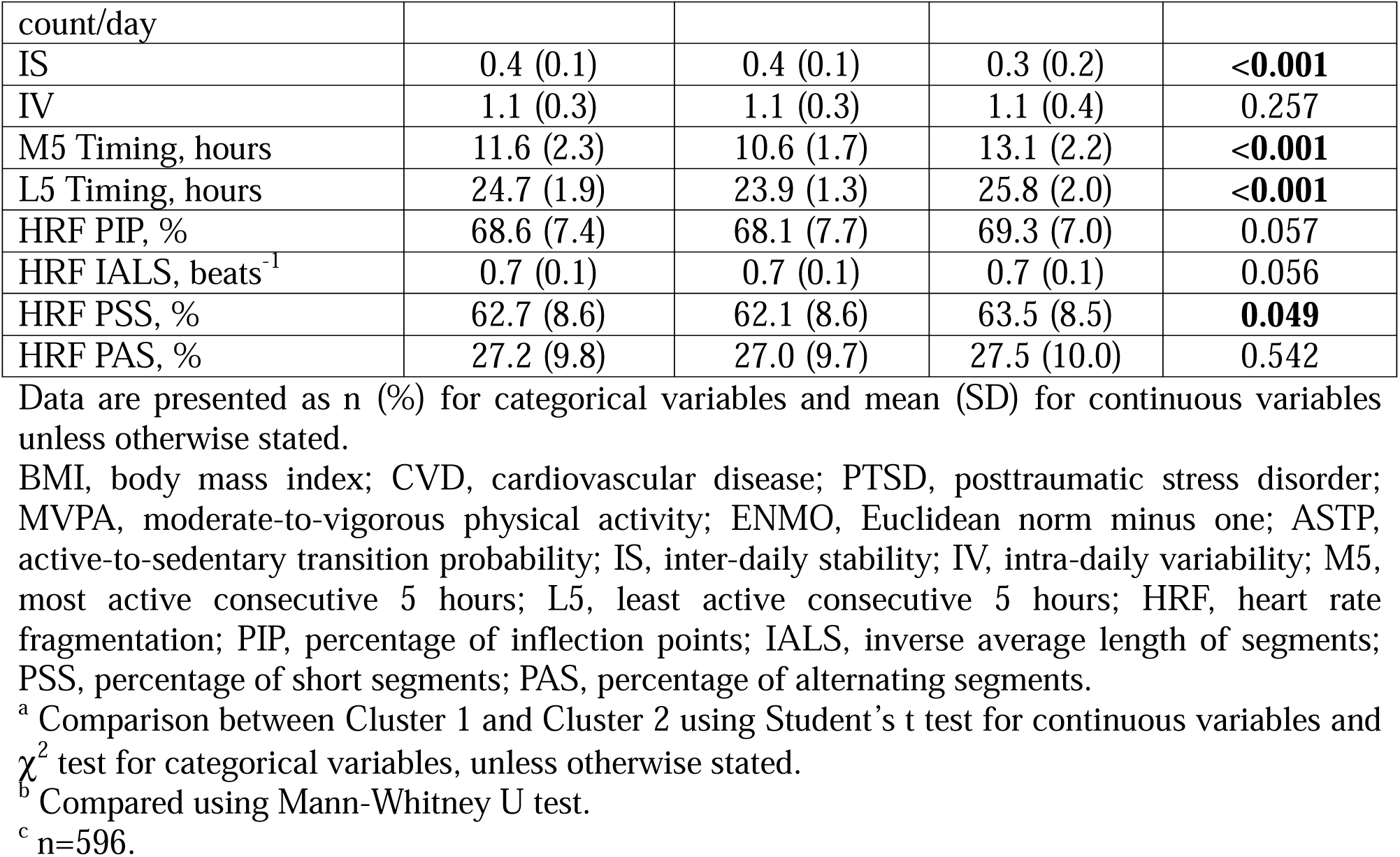
Baseline Characteristics Overall and by Digital Phenotype of Physical Activity Patterns.

|  |  | **Total**<br>**(n=602)** | **Cluster 1**<br>**(n=341)** | **Cluster 2**<br>**(n=261)** | **P-value<sup>a</sup>** |
| --- | --- | --- | --- | --- | --- |
| Age, years |  | 73.8 (2.9) | 73.9 (3.0) | 73.6 (2.9) | 0.171 |
| Race, n (%) |  |  |  |  | 0.922 |
| White |  | 561 (93.2) | 317 (93.0) | 244 (93.5) |  |
| Black |  | 30 (5.0) | 18 (5.3) | 12 (4.6) |  |
| Other |  | 11 (1.8) | 6 (1.8) | 5 (1.9) |  |
| Education, n (%) |  |  |  |  | 0.550 |
| Elementary/Middle School |  | 3 (0.5) | 3 (0.9) | 0 (0.0) |  |
| High School |  | 116 (19.3) | 70 (20.5) | 46 (17.6) |  |
| Vocational/Technical School |  | 54 (9.0) | 32 (9.4) | 22 (8.4) |  |
| Some College |  | 192 (31.9) | 100 (29.3) | 92 (35.3) |  |
| College Degree |  | 181 (30.1) | 104 (30.5) | 77 (29.5) |  |
| Post-Graduate College |  | 39 (6.5) | 23 (6.7) | 16 (6.1) |  |
| Advanced Degree |  | 17 (2.8) | 9 (2.6) | 8 (3.1) |  |
| Smoking Status, n (%) |  |  |  |  | 0.188 |
| Current Smoker |  | 43 (7.1) | 19 (5.6) | 24 (9.2) |  |
| Former Smoker |  | 319 (53.0) | 180 (52.8) | 139 (53.3) |  |
| Never Smoker |  | 240 (39.9) | 142 (41.6) | 98 (37.6) |  |
| Drinking Status, days | mean (SD) | 5.7 (9.4) | 6.3 (9.9) | 4.9 (8.7) | 0.069 |
|  | median (IQR) | 0.0 (6.0) | 1.0 (9.0) | 0.0 (5.0) | 0.122<sup>b</sup> |
| BMI, kg/m<sup>2</sup> |  | 28.5 (4.8) | 28.4 (4.8) | 28.7 (4.8) | 0.490 |
| Hypertension, n (%) |  | 407 (67.6) | 222 (65.1) | 185 (70.9) | 0.133 |
| Diabetes, n (%) |  | 163 (27.1) | 79 (23.2) | 84 (32.2) | **0.014** |
| CVD, n (%) |  | 202 (33.6) | 110 (32.3) | 92 (35.3) | 0.441 |
| Depression<sup>c</sup>, n (%) |  | 113 (19.0) | 51 (15.0) | 62 (24.2) | **0.005** |
| PTSD<sup>c</sup>, n (%) |  | 90 (15.1) | 50 (14.7) | 40 (15.6) | 0.756 |
| Step Count, steps/day |  | 6908.9 (2776.0) | 7255.6 (2040.4) | 6456.0 (3464.9) | **0.001** |
| Time Spent Sedentary, % |  | 72.6 (6.9) | 71.7 (5.7) | 73.9 (8.0) | **<0.001** |
| Time Spent MVPA, % |  | 8.2 (3.9) | 8.7 (3.0) | 7.7 (4.8) | **0.002** |
| ENMO, mg | mean (SD) | 17.3 (10.5) | 17.7 (10.6) | 16.8 (10.3) | 0.292 |
|  | median (IQR) | 16.2 (6.8) | 16.9 (5.6) | 15.2 (9.3) | **<0.001<sup>b</sup>** |
| Intensity Gradient |  | -3.0 (0.2) | -3.0 (0.2) | -3.0 (0.2) | **<0.001** |
| ASTP, % |  | 25.7 (8.4) | 24.1 (6.0) | 27.7 (10.6) | **<0.001** |
| Inactive Bout Length, minutes | mean (SD) | 43.4 (45.3) | 32.2 (18.5) | 58.0 (62.6) | **<0.001** |
|  | median (IQR) | 30.2 (26.9) | 28.7 (17.9) | 35.8 (51.2) | **<0.001<sup>b</sup>** |
| Sedentary Bouts ≥ 30min, |  | 7.3 (1.7) | 7.9 (1.4) | 6.5 (1.8) | **<0.001** |

|  |  |  |  |  |
| --- | --- | --- | --- | --- |
| count/day |  |  |  |  |
| IS | 0.4 (0.1) | 0.4 (0.1) | 0.3 (0.2) | **<0.001** |
| IV | 1.1 (0.3) | 1.1 (0.3) | 1.1 (0.4) | 0.257 |
| M5 Timing, hours | 11.6 (2.3) | 10.6 (1.7) | 13.1 (2.2) | **<0.001** |
| L5 Timing, hours | 24.7 (1.9) | 23.9 (1.3) | 25.8 (2.0) | **<0.001** |
| HRF PIP, % | 68.6 (7.4) | 68.1 (7.7) | 69.3 (7.0) | 0.057 |
| HRF IALS, beats<sup>-1</sup> | 0.7 (0.1) | 0.7 (0.1) | 0.7 (0.1) | 0.056 |
| HRF PSS, % | 62.7 (8.6) | 62.1 (8.6) | 63.5 (8.5) | **0.049** |
| HRF PAS, % | 27.2 (9.8) | 27.0 (9.7) | 27.5 (10.0) | 0.542 |

Data are presented as n (%) for categorical variables and mean (SD) for continuous variables unless otherwise stated.
BMI, body mass index; CVD, cardiovascular disease; PTSD, posttraumatic stress disorder; MVPA, moderate-to-vigorous physical activity; ENMO, Euclidean norm minus one; ASTP, active-to-sedentary transition probability; IS, inter-daily stability; IV, intra-daily variability; M5, most active consecutive 5 hours; L5, least active consecutive 5 hours; HRF, heart rate fragmentation; PIP, percentage of inflection points; IALS, inverse average length of segments; PSS, percentage of short segments; PAS, percentage of alternating segments.
<sup>a</sup> Comparison between Cluster 1 and Cluster 2 using Student's t test for continuous variables and $\chi^2$ test for categorical variables, unless otherwise stated.
<sup>b</sup> Compared using Mann-Whitney U test.
<sup>c</sup> n=596.

We performed several sensitivity analyses to address the robustness of our findings. First, covariates such as hypertension and diabetes were evaluated in sensitivity analyses further adjusting for them, as their role as confounders or mediators cannot be definitively established. To address sparsity of the lowest education category, additional sensitivity analyses evaluated model stability after excluding the 3 participants in the lowest education category. Finally, to evaluate whether the multi-dimensional digital phenotype of physical activity patterns provided added value beyond individual principal component scores, the retained principal components and respective interaction terms were evaluated for associations with heart rate fragmentation. These continuous principal component score models with interaction terms were then compared with the phenotype models regarding model fit, as assessed by Akaike Information Criterion (AIC).

All statistical analyses were performed using SAS software (Version 9.4; SAS Institute, Cary, NC, US) and R (Version 4.5.1). Principal component analysis and Horn’s parallel analysis were conducted using the psych package,^34^ and Gaussian mixture models were fitted using the mclust package.^35^ A two-sided p-value < 0.05 was considered statistically significant.

## Results

### Unsupervised Machine Learning Phenotyping

From Horn’s parallel analysis, the first two components were retained (Fiue S1). The two components accounted for 60% of the total variance in physical activity, with the first and second components explaining 46% and 14%, respectively (Table S1). The first principal component exhibited strong positive loading values for step count (0.90), time spent MVPA (0.89), and ENMO (0.84), and strong negative loading values for inactive bout length (-0.8), time spent sedentary (-0.83), and ASTP (-0.80 (Tabe S1). The second principal component showed strong positive loading values for M5 timing (0.86) and L5 timing (0.86) (Table S1).

The dendrogram derived from agglomerative hierarchical clustering established candidate cluster values ranging from k=2 to 5 (Figure S2). Subsequent evaluation of optimal cluster value using Gaussian mixture models based on BIC identified an optimal value of k=2 (Table S2). The resulting Gaussian mixture model with k=2 yielded mean and median assignment probabilities of 64.68% and 62.86%, respectively (Tabe S3). Because the minimum possible assignment probability for a two-cluster model is 50%, these values indicate substantial overlap between clusters and considerable classification uncertainty; one quarter of participants had assignment probabilities below 57.18% (Table S3). From the cluster assignment plot, separation between the two clusters was non-linear (Figure 2). Cluster 1 formed a compact group at intermediate first principal component scores and predominantly lower second principal component scores (earlier timing), whereas cluster 2 formed a diffuse, crescent-shaped distribution surrounding cluster 1, concentrated at higher second principal component scores (later timing) and extending to both extremes of the first principal component (i.e., including both the least and most active participants). Cluster membership therefore did not vary monotonically along either component, and the two cluster centroids were located close together. Cluster 1 and cluster 2 are hereafter referred to as the compact-early and diffuse-late phenotypes, respectively (VII spherical, varying volume model; cluster 1 mean principal component 1 score: 0.057, principal component 2 score: -0.288, variance: 0.686; cluster 2 mean principal component 1 score: -0.03, principal component 2: 0.267, variance: 1.211; Figure 2). The standardized z-score heatmap showed that the phenotypes differed most in diurnal timing and prolonged sedentary bouts: compared with the diffuse-late phenotype, the compact-early phenotype had earlier M5 and L5 timing (mean z-scores −0.47 and −0.44 vs. 0.62 and 0.57) and more sedentary bouts ≥ 30 minutes (0.35 vs. −0.45). Differences in volume, intensity, fragmentation, and variability features were smaller, with the compact-early phenotype showing slightly greater step count, time spent MVPA, IG, ENMO, and IS, and slightly lower time spent sedentary, ASTP, and inactive bout length; IV was similar between phenotypes (Figure 3). To address potential non-independence of co-twins, the same unsupervised machine learning phenotyping pipeline was performed on 314 independent participants whose co-twins were not present in the dataset, yielding consistent findings (Tabl S4).

**Figure 2.**
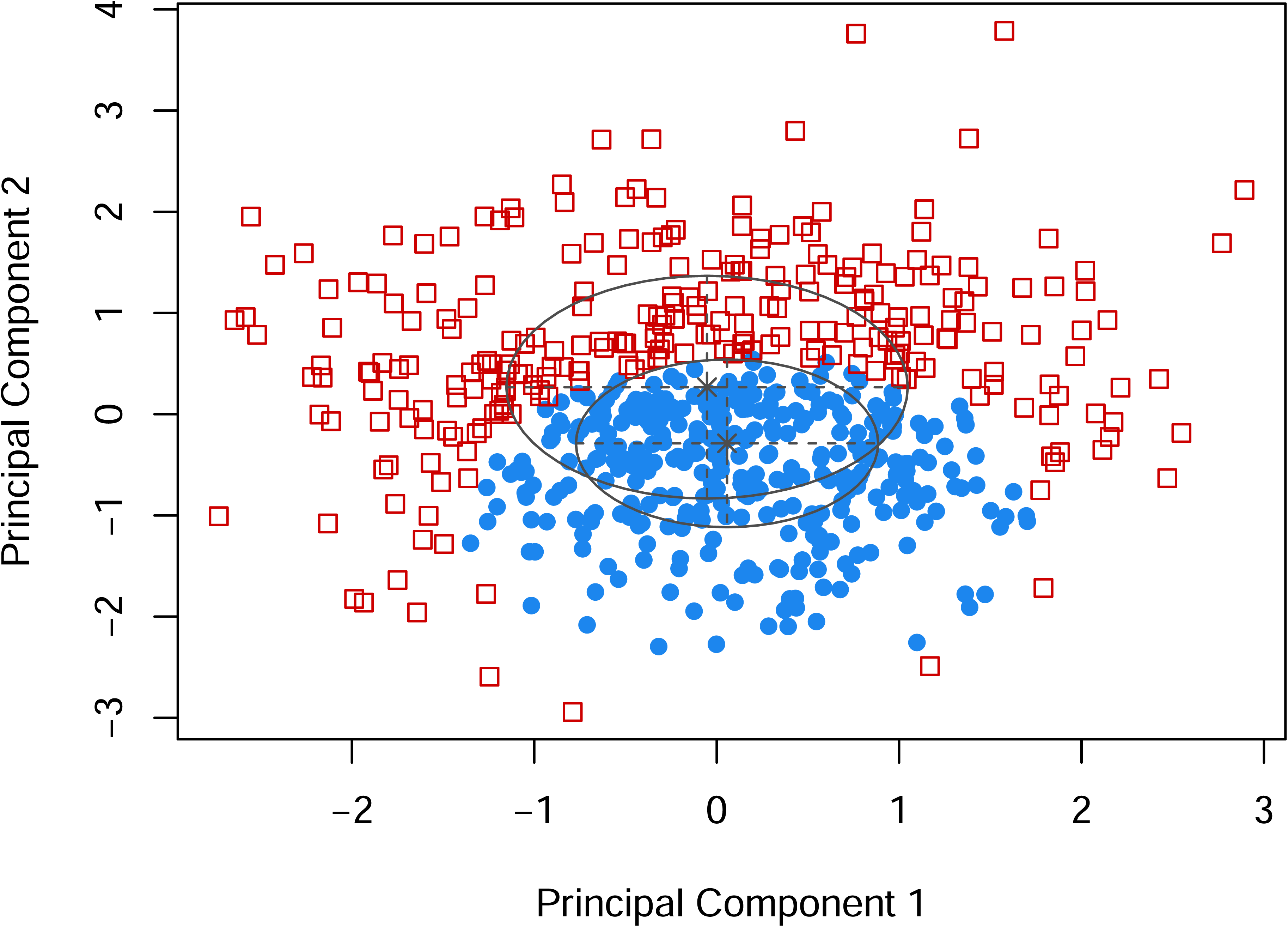
Cluster assignment plot based on the two retained principal components, derived from Gaussian mixture model with k=2. Blue filled circles and red open squares represent cluster 1 (compact-early phenotype) (n=341) and cluster 2 (diffuse-late phenotype) (n=261), respectively. Ellipses and asterisks indicate the estimated covariance and mean of each mixture component. (VII spherical, varying volume model; cluster 1 mean principal component 1 score: 0.057, principal component 2 score: -0.288, variance: 0.686; cluster 2 mean principal component 1 score: -0053, principal component 2: 0.267, variance: 1.211). The first principal component reflects overall activity volume, and higher second principal component scores indicate later activity timing.

**Figure 3.**
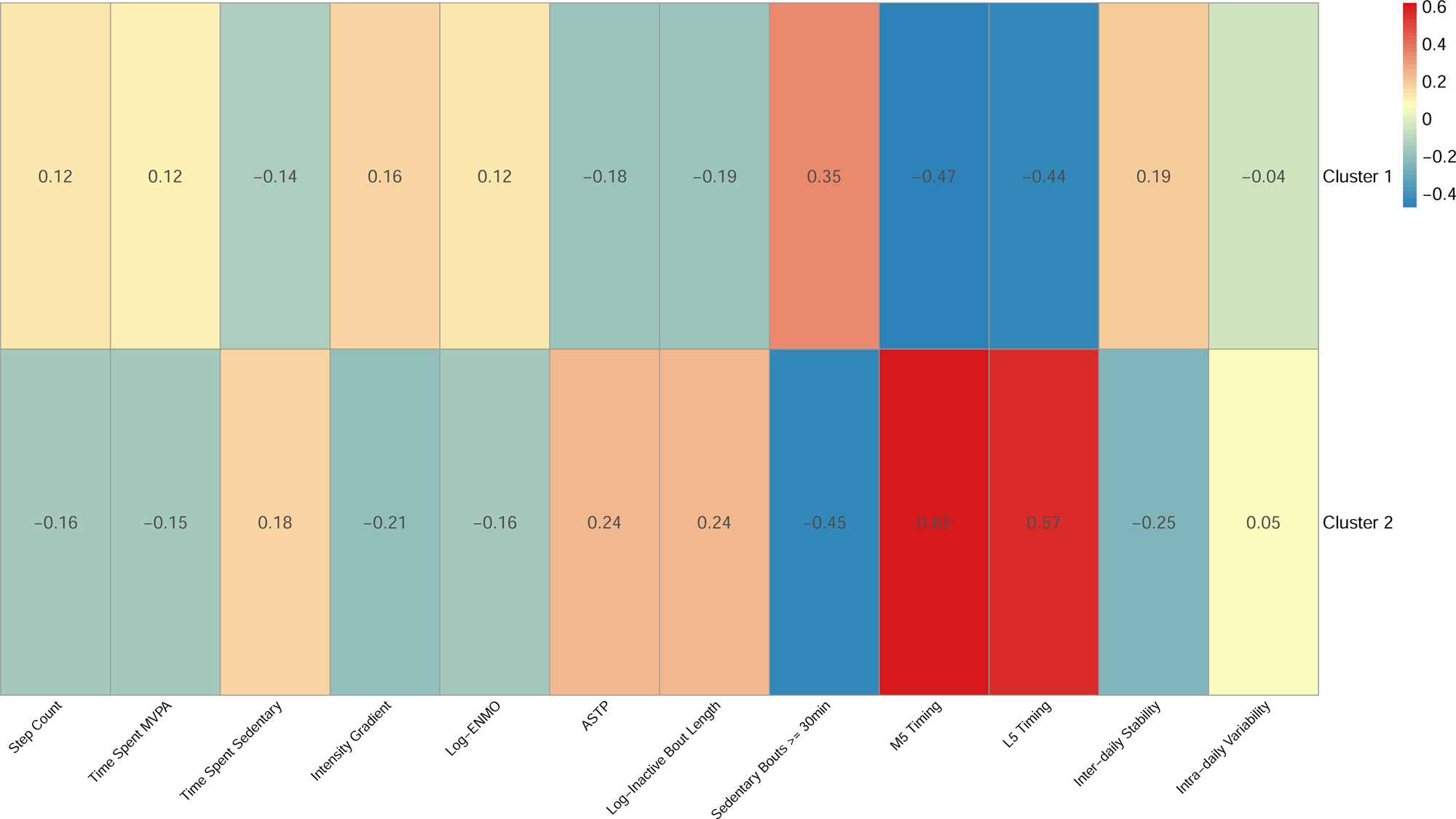
Standardized z-score heatmap of mean feature profiles by cluster 1 (compact-early phenotype) (n=341) and cluster 2 (diffuse-late phenotype) (n=261). Values represent mean z-scores. MVPA, moderate-to-vigorous physical activity; ENMO, Euclidean norm minus one; IG, intensity gradient; IS, inter-daily stability; IV, intra-daily variability; ASTP, active-to-sedentary transition probability; M5, most active consecutive 5 hours; L5, least active consecutive 5 hours.

### General Characteristics

Of the 602 participants included in the analysis, the mean age was 73.8 ± 2.9 years and 93.2% were White participants (Table 1). Included participants were younger (73.8 ± 2.9 years vs. 74.8 ± 3.4 years) and had a lower prevalence of post-graduate education than participants excluded from the analysis (Table S5).

Table 1 shows participant demographics stratified according to respective digital phenotypes of physical activity patterns. Compared with the diffuse-late phenotype, participants in the compact-early phenotype had a lower prevalence of diabetes and lifetime depression. They also had greater step count, time spent MVPA, median ENMO, IG, sedentary bouts ≥ 30 minutes, and IS, as well as lower time spent sedentary, ASTP, inactive bout length, PSS, M5 timing, and L5 timing. PIP, IALS, and PAS were slightly higher in the diffuse-late phenotype, but these differences did not reach statistical significance.

When stratifying the 602 participants into tertiles of the first principal component, the prevalence of current smoking, hypertension, and diabetes differed across tertiles, with the lowest prevalence in the highest tertile (Tale S6). BMI, PIP, and IALS also differed across tertiles, with the lowest values in the highest tertile (Table S6).

Similarly, when stratifying the 602 participants into tertiles of the second principal component, the prevalence of diabetes differed across tertiles, with the highest prevalence in the highest tertile (Tale S7).

### Associations between Digital Phenotype of Physical Activity and Heart Rate Fragmentation

In fully adjusted models, the diffuse-late phenotype was associated with higher PIP, the primary outcome (β: 1.199, 95% CI: 0.034 to 2.363, p-value: 0.044), a difference of approximately 0.16 SD. Among secondary outcomes, the diffuse-late phenotype was also associated with higher IALS (β: 0.01210, 95% CI: 0.00048 to 0.02373, p-value: 0.041) compared with the compact-early phenotype (reference) (Table 2). There were no significant associations between digital phenotypes of physical activity patterns and PSS or PAS (Table 2).

**Table 2.** Association between Digital Phenotype of Physical Activity Patterns and Heart Rate Fragmentation.

|  | Model | Cluster 2 (Diffuse-Late Phenotype) |  |
| --- | --- | --- | --- |
| | | $\beta$ (95% CI) | P-value |
| HRF PIP, % | Model 1<sup>a</sup> | 1.171 (-0.037, 2.380) | 0.057 |
|  | Model 2<sup>b</sup> | 1.266 (0.068, 2.463) | **0.038** |
|  | Model 3<sup>c</sup> | 1.199 (0.034, 2.363) | **0.044** |
| HRF IALS, beats<sup>-1</sup> | Model 1<sup>a</sup> | 0.01174 (-0.00032, 0.02380) | 0.056 |
|  | Model 2<sup>b</sup> | 0.01277 (0.00081, 0.02474) | **0.037** |
|  | Model 3<sup>c</sup> | 0.01210 (0.00048, 0.02373) | **0.041** |
| HRF PSS, % | Model 1<sup>a</sup> | 1.274 (-0.115, 2.662) | 0.072 |
|  | Model 2<sup>b</sup> | 1.055 (-0.304, 2.413) | 0.127 |
|  | Model 3<sup>c</sup> | 1.017 (-0.323, 2.356) | 0.136 |
| HRF PAS, % | Model 1<sup>a</sup> | 0.356 (-1.235, 1.947) | 0.659 |
|  | Model 2<sup>b</sup> | 0.121 (-1.452, 1.695) | 0.879 |
|  | Model 3<sup>c</sup> | 0.103 (-1.463, 1.670) | 0.896 |

Each heart rate fragmentation outcome was tested separately, with Cluster 1 serving as the reference group.
CI, confidence interval; HRF, heart rate fragmentation; PIP, percentage of inflection points; IALS, inverse average length of segments; PSS, percentage of short segments; PAS, percentage of alternating segments.
<sup>a</sup> Crude model with twin ID as random intercept.
<sup>b</sup> Adjusted for age, race, education, BMI, smoking, and drinking, with twin ID as random intercept.
<sup>c</sup> Adjusted for age, race, education, BMI, smoking, drinking, and history of any composite CVD, with twin ID as random intercept.

Sensitivity analyses conducted to assess model stability against sparsity of the lowest education category yielded similar results for PIP (β: 1.187, 95% CI: 0.025 to 2.349, p-value: 0.045) and IALS (β: 0.01198, 95% CI: 0.00038 to 0.02359, p-value: 0.043) in fully adjusted models (Table S8). No significant associations were observed for PSS or PAS (Tale S8).

When additionally adjusting for hypertension, the diffuse-late phenotype was associated with higher PIP (β: 1.218, 95% CI: 0.052 to 2.384, p-value: 0.041) and higher IALS (β: 0.01228, 95% CI: 0.00064 to 0.02392, p-value: 0.039) compared with the compact-early phenotype (Table 3). However, these associations were attenuated and lost significance when additionally adjusting for diabetes or both hypertension and diabetes (Table 3). No significant associations were observed for PSS or PAS (Table 3).

**Table 3.** Sensitivity Analysis of the Association between Digital Phenotype of Physical Activity Patterns and Heart Rate Fragmentation With Additional Adjustment for Diabetes and Hypertension.

|  | Model | Cluster 2 (Diffuse-Late Phenotype) |  |
| --- | --- | --- | --- |
| | | $\beta$ (95% CI) | *P*-value |
| HRF PIP, % | Model 4<sup>a</sup> | 1.218 (0.052, 2.384) | **0.041** |
|  | Model 5<sup>b</sup> | 0.993 (-0.161, 2.146) | 0.091 |
|  | Model 6<sup>c</sup> | 1.016 (-0.139, 2.170) | 0.084 |
| HRF IALS, beats<sup>-1</sup> | Model 4<sup>a</sup> | 0.01228 (0.00064, 0.02392) | **0.039** |
|  | Model 5<sup>b</sup> | 0.01012 (-0.00141, 0.02165) | 0.085 |
|  | Model 6<sup>c</sup> | 0.01034 (-0.00120, 0.02187) | 0.079 |
| HRF PSS, % | Model 4<sup>a</sup> | 0.964 (-0.373, 2.300) | 0.156 |
|  | Model 5<sup>b</sup> | 0.926 (-0.415, 2.267) | 0.174 |
|  | Model 6<sup>c</sup> | 0.884 (-0.455, 2.222) | 0.194 |
| HRF PAS, % | Model 4<sup>a</sup> | 0.037 (-1.525, 1.600) | 0.962 |
|  | Model 5<sup>b</sup> | 0.124 (-1.448, 1.697) | 0.876 |
|  | Model 6<sup>c</sup> | 0.070 (-1.498, 1.638) | 0.930 |

Each heart rate fragmentation outcome was tested separately, with Cluster 1 serving as the reference group.
CI, confidence interval; HRF, heart rate fragmentation; PIP, percentage of inflection points; IALS, inverse average length of segments; PSS, percentage of short segments; PAS, percentage of alternating segments.
<sup>a</sup> Adjusted for Model 3 covariates and hypertension, with twin ID as random intercept.
<sup>b</sup> Adjusted for Model 3 covariates and diabetes, with twin ID as random intercept.
<sup>c</sup> Adjusted for Model 3 covariates, hypertension, and diabetes, with twin ID as random intercept.

In fully adjusted models evaluating the associations of principal component scores and respective interaction terms with heart rate fragmentation, neither the main effects nor the interaction terms were significantly associated with any measure of heart rate fragmentation (Table S9). Furthermore, compared with the principal component score models with interaction terms, the phenotype model consistently had better model fit, as assessed by AIC, across all heart rate fragmentation metrics (Tale S10).

## Discussion

In this cross-sectional study of older male veterans, we identified two overlapping phenotypes of physical activity based on different clustering patterns of principal components focused on physical activity and diurnal activity patterns. The compact-early phenotype formed a relatively homogeneous group characterized by earlier activity timing and slightly greater overall activity, whereas the diffuse-late phenotype was more heterogeneous, characterized primarily by later activity timing and spanning both low and high activity volume. The diffuse-late phenotype was associated with modestly higher PIP, the primary outcome, and higher IALS compared with the compact-early phenotype. However, no significant associations between digital phenotypes of physical activity patterns and PSS or PAS were observed. These findings parallel prior work linking physical activity with more favorable cardiac autonomic function, although direct comparisons are limited because heart rate fragmentation captures properties distinct from heart rate variability and prior studies largely evaluated exercise interventions rather than habitual activity patterns.^8^ While such studies have largely focused on differences in traditional modalities of physical activity (aerobic vs. resistance),^8^ the current study highlights the associations between multi-dimensional digital phenotypes of physical activity patterns and heart rate fragmentation, and the value of evaluating holistic, multi-dimensional phenotypes of physical activity patterns. As AIC comparisons demonstrated that the phenotype models consistently yielded better model fit compared to continuous principal component score models incorporating interaction terms across all heart rate fragmentation measures, phenotyping physical activity patterns may provide meaningful value beyond standard continuous principal component scores. Notably, the phenotypes differed most along the diurnal timing component, suggesting that when activity occurs, and not only how much, may be relevant to cardiac rhythm regulation, although heart rate fragmentation did not differ across tertiles of the timing component alone; activity timing has rarely been examined in relation to cardiac autonomic or sinoatrial function.

This study has several potential implications. As PIP and IALS may represent the overall fragmentation by capturing the frequency and density of directional changes in cardiac inter-beat intervals,^5^ while PSS and PAS may represent specialized patterns of fragmentation by capturing the accumulation of short and alternating runs of heart rate acceleration and deceleration,^5^ our findings suggest that the diffuse-late phenotype may relate more to overall fragmentation than to specific fragmentation patterns; however, associations with PSS and PAS were not statistically significant, and the PSS estimate was similar in direction to that for PIP. Furthermore, the modest attenuation in magnitude of estimates when further adjusting for diabetes suggests that metabolic and glycemic regulation may partly explain the association between the digital phenotype of physical activity patterns and heart rate fragmentation.^36, 37^ This potential physiological mechanism is further suggested by the underlying structure of the multi-dimensional digital phenotypes of physical activity patterns. While higher first principal component scores were accompanied by a more favorable cardiometabolic and lifestyle profile, including a lower prevalence of current smoking, hypertension, and diabetes and lower BMI, the second component was defined by diurnal patterns, such as M5 and L5 timings, and higher scores were accompanied by a higher prevalence of diabetes. Considering the role of diurnal patterns as a proxy for circadian rhythm,^38^ and the associations of circadian rhythm with metabolic and glycemic regulation,^39^ the digital phenotypes of physical activity patterns may represent the manifestation of heart rate fragmentation risk profiles at the intersection of general health, lifestyle, and circadian-metabolic regulation, though this speculative mechanism warrants future studies for confirmation. Together, these findings may help identify specific phenotypes of physical activity patterns that could be promoted for the prevention of CVD and targeted for personalized exercise interventions, particularly in aging populations.

The current study has several strengths, including the objectively measured physical activity parameters, coverage of multiple physical activity domains, continuous ambulatory electrocardiogram measurement, derivation of multi-dimensional phenotypes of physical activity patterns, and incorporation of multiple covariates. Despite these strengths, we acknowledge several limitations. The observational nature of the study limits causal inference from these findings. Furthermore, the cross-sectional study design prevents us from ruling out reverse causality, whereby participants with lower heart rate fragmentation or the absence of underlying diseases are more likely to be part of the compact-early phenotype. Included participants were younger and less likely to have post-graduate education than excluded participants, although other lifestyle and clinical variables did not differ significantly; in addition, heart rate fragmentation data were missing for approximately 18% of the cohort because processing was incomplete at the time of this interim analysis, so selection bias cannot be ruled out. The Gaussian mixture model showed considerable classification uncertainty (mean assignment probability 64.68%, where 50% is the minimum for two clusters), and the cluster solution has not been internally or externally validated; such misclassification would generally be expected to bias associations toward the null, although this was not formally assessed. The association with PIP was small (approximately 0.16 SD), and secondary outcomes were not corrected for multiple comparisons. Medications affecting heart rate dynamics (e.g. beta-blockers, non-dihydropyridine calcium-channel blockers, antiarrhythmics, and antidepressants) were not included in the models, and residual confounding by these factors is possible. Heart rate fragmentation was computed across the full recording, including periods of activity and rest, so phenotype differences may partly reflect when and how much participants were active during monitoring, and ectopic beats were removed only through an RR-interval change filter rather than explicit beat classification. In addition, the Freedson cut points were calibrated for hip-worn devices; despite enabling the “Worn on Wrist” setting, applying them to wrist-derived, waist-equivalent counts may misclassify sedentary time and MVPA in older adults, and wrist-worn devices cannot distinguish sitting, reclining, and standing still. The population used for this analysis was exclusively male and predominantly of older age and White race; therefore, the generalizability of these findings to other populations may be limited. Finally, despite the incorporation of multiple covariates, the possibility of unknown confounding persists.

## Conclusion

In this cross-sectional study of older male veterans, we observed two new types of findings. First, principal component analysis of 12 accelerometry-derived features identified two components representing overall activity volume and diurnal activity timing. Second, Gaussian mixture modeling of these components identified a focal, compact-early phenotype and a diffuse-late phenotype that surrounded it, reflecting different mixtures of the two components. The diffuse-late phenotype exhibited modestly higher PIP, the primary outcome, and higher IALS compared with the compact-early phenotype. However, there were no significant associations between digital phenotypes of physical activity patterns and PSS or PAS. These heart rate fragmentation differences highlight potential significance for CVD that should be further explored in future research, including whether such phenotypes could inform CVD prevention and personalized exercise interventions. Future studies should aim to validate such digital phenotypes across diverse populations and investigate their longitudinal associations with heart rate fragmentation.

## Data Availability

Data are available from the Vietnam Era Twins Study of Electrophysiology in Posttraumatic Stress Disorder (VETS-EP) Study upon reasonable request to the corresponding author; restrictions apply and data are not publicly available

## Acknowledgements

The authors thank the staff and participants of the VETS-EP study for their important contributions.

## Supplemental Material

Tables S1-S10, Figres S1-S2

