## Supplemental Material for "A Novel Digital Phenotype of Physical Activity: Associations with Heart Rate Fragmentation"

**Table S1.** Principal Component Analysis Eigenvalues, Loadings, Communalities, and Variance Explained for the 12 Physical Activity Variables

|  | **PC1 (EV=5.52)** | **PC2 (EV=1.67)** | **Communality** |
| --- | --- | --- | --- |
| Step Count, steps/day | 0.90 | -0.12 | 0.82 |
| Time Spent MVPA, % | 0.89 | -0.11 | 0.80 |
| Inactive Bout Length^a^, minutes | -0.86 | 0.09 | 0.75 |
| ENMO^a^, mg | 0.84 | -0.01 | 0.70 |
| Time Spent Sedentary, % | -0.83 | 0.07 | 0.69 |
| ASTP, % | -0.80 | 0.10 | 0.64 |
| Intensity Gradient | 0.63 | -0.16 | 0.42 |
| IV | -0.60 | -0.07 | 0.36 |
| IS | 0.59 | -0.24 | 0.41 |
| M5 Timing, hours | -0.07 | 0.86 | 0.74 |
| L5 Timing, hours | 0.05 | 0.86 | 0.74 |
| Sedentary Bouts ≥ 30min, count/day | 0.22 | -0.24 | 0.11 |
| Variance Explained (%) | 46 | 14 | - |
| Cumulative Variance (%) | 46 | 60 | - |

Varimax rotation was applied, and 2 components were extracted.

PC, principal component; EV, eigenvalue; MVPA, moderate-to-vigorous physical activity; ENMO, Euclidean norm minus one; ASTP, active-to-sedentary transition probability; IS, inter-daily stability; IV, intra-daily variability; M5, most active consecutive 5 hours; L5, least active consecutive 5 hours.

^a^ Log-transformations were applied due to skewness.

**Table S2.** Comparison of Bayesian Information Criterion Values for Gaussian Mixture Models with Varying Numbers of Clusters

|  | **BIC** |
| --- | --- |
| 2 | **-3450.26** |
| 3 | -3458.20 |
| 4 | -3476.84 |
| 5 | -3495.60 |

BIC values were obtained from the Gaussian mixture model implemented in the mclust package; larger BIC values indicate better model fit.

BIC, Bayesian information criterion.

|  | Value |
| --- | --- |
| Minimum Assignment Probability, % | 50.08 |
| 1^st^ Quartile Assignment Probability, % | 57.18 |
| Median Assignment Probability, % | 62.86 |
| Mean Assignment Probability, % | 64.68 |
| 3^rd^ Quartile Assignment Probability, % | 67.77 |
| Maximum Assignment Probability, % | 99.92 |

**Table S3.** Gaussian Mixture Model Classification Metrics (k=2)

Assignment probabilities represent the maximum posterior probability per participant.

GMM, Gaussian mixture model.

|  | Parameter | Value |
| --- | --- | --- |
| PCA | Components Retained | 2 |
|  | EV (PC1) | 5.57 |
|  | EV (PC2) | 1.83 |
|  | Variance explained (PC1) | 46% |
|  | Variance explained (PC2) | 15% |
| GMM | Optimal k | 2 |
|  | Selected Covariance Parameterization | VII (spherical, varying volume) |

**Table S4.** Summary of Unsupervised Machine Learning Phenotyping Pipeline among 314 Independent Participants

PCA, principal component analysis; EV, eigenvalue; GMM, Gaussian mixture model.

**Table S5.** Comparison of Baseline Characteristics between Included and Excluded Participants

|  | | **Included**  **(n=602)** | **Excluded**  **(n=341)** | ***P-*value^a^** |
| --- | --- | --- | --- | --- |
| Age, years | | 73.8 (2.9) | 74.8 (3.4) | **<0.001** |
| Race, n (%) | |  |  | 0.936 |
| White | | 561 (93.2) | 307 (93.3) |  |
| Black | | 30 (5.0) | 17 (5.2) |  |
| Other | | 11 (1.8) | 5 (1.5) |  |
| Education, n (%) | |  |  | **0.039** |
| Elementary/Middle School | | 3 (0.5) | 1 (0.3) |  |
| High School | | 116 (19.3) | 52 (15.3) |  |
| Vocational/Technical School | | 54 (9.0) | 32 (9.4) |  |
| Some College | | 192 (31.9) | 101 (29.7) |  |
| College Degree | | 181 (30.1) | 98 (28.8) |  |
| Post-Graduate College | | 39 (6.5) | 44 (12.9) |  |
| Advanced Degree | | 17 (2.8) | 12 (3.5) |  |
| Smoking Status, n (%) | |  |  | 0.320 |
| Current Smoker | | 43 (7.1) | 20 (6.2) |  |
| Former Smoker | | 319 (53.0) | 159 (48.9) |  |
| Never Smoker | | 240 (39.9) | 146 (44.9) |  |
| Drinking Status, days | mean (SD) | 5.7 (9.4) | 5.6 (9.0) | 0.911 |
|  | median (IQR) | 0.0 (6.0) | 0.0 (8.0) | 0.740^b^ |
| BMI, kg/m^2^ | | 28.5 (4.8) | 28.6 (4.8) | 0.796 |
| Hypertension, n (%) | | 407 (67.6) | 200 (61.5) | 0.064 |
| Diabetes, n (%) | | 163 (27.1) | 103 (31.7) | 0.138 |
| CVD, n (%) | | 202 (33.6) | 97 (29.9) | 0.249 |

Data are presented as n (%) for categorical variables and mean (SD) for continuous variables unless otherwise stated.

Sample sizes for excluded participants vary across characteristics due to missing data.

BMI, body mass index; CVD, cardiovascular disease.

^a^ Comparison between included and excluded participants using Student’s t test for continuous variables and χ^2^ test for categorical variables, unless otherwise stated.

^b^ Compared using Mann-Whitney U test.

**Table S6**. Comparison of Baseline Characteristics between Tertiles of Principal Component 1

|  | | **Low Tertile**  **(< -0.4)**  **(n=200)** | **Middle Tertile**  **(-0.4 to 0.5)**  **(n=201)** | **High Tertile**  **(> 0.5)**  **(n=201)** | ***P-*value^a^** |
| --- | --- | --- | --- | --- | --- |
| PC 1, score | | -1.1 (0.5) | 0.0 (0.2) | 1.1 (0.5) | **<0.001** |
| Age, years | | 74.1 (2.9) | 73.6 (3.1) | 73.6 (2.8) | 0.108 |
| Race, n (%) | |  |  |  | 0.261 |
| White | | 185 (92.5) | 184 (91.5) | 192 (95.5) |  |
| Black | | 13 (6.5) | 12 (6.0) | 5 (2.5) |  |
| Other | | 2 (1.0) | 5 (2.5) | 4 (2.0) |  |
| Education, n (%) | |  |  |  | 0.506 |
| Elementary/Middle School | | 1 (0.5) | 1 (0.5) | 1 (0.5) |  |
| High School | | 43 (21.5) | 37 (18.4) | 36 (17.9) |  |
| Vocational/Technical School | | 20 (10.0) | 14 (7.0) | 20 (10.0) |  |
| Some College | | 68 (34.0) | 71 (35.3) | 53 (26.4) |  |
| College Degree | | 49 (24.5) | 58 (28.9) | 74 (36.8) |  |
| Post-Graduate College | | 12 (6.0) | 14 (7.0) | 13 (6.5) |  |
| Advanced Degree | | 7 (3.5) | 6 (3.0) | 4 (2.0) |  |
| Smoking Status, n (%) | |  |  |  | **0.010** |
| Current Smoker | | 23 (11.5) | 13 (6.5) | 7 (3.5) |  |
| Former Smoker | | 111 (55.5) | 104 (51.7) | 104 (51.7) |  |
| Never Smoker | | 66 (33.0) | 84 (41.8) | 90 (44.8) |  |
| Drinking Status, days | mean (SD) | 4.2 (8.1) | 6.1 (9.8) | 6.9 (10.2) | **0.015** |
|  | median (IQR) | 0 (4.5) | 1 (6.0) | 1 (10.0) | 0.058^b^ |
| BMI, kg/m^2^ | | 30.0 (5.7) | 28.4 (4.3) | 27.1 (3.8) | **<0.001** |
| Hypertension, n (%) | | 144 (72.0) | 142 (70.7) | 121 (60.2) | **0.022** |
| Diabetes, n (%) | | 77 (38.5) | 53 (26.4) | 33 (16.4) | **<0.001** |
| CVD, n (%) | | 76 (38.0) | 71 (35.3) | 55 (27.4) | 0.064 |
| HRF PIP, % | | 69.7 (7.7) | 68.4 (6.9) | 67.8 (7.6) | **0.038** |
| HRF IALS, beats^-1^ | | 0.7 (0.1) | 0.7 (0.1) | 0.7 (0.1) | **0.049** |
| HRF PSS, % | | 64.8 (9.3) | 61.3 (7.4) | 61.9 (8.7) | **<0.001** |
| HRF PAS, % | | 29.1 (10.8) | 25.5 (8.5) | 27.1 (9.7) | **0.001** |

Data are presented as n (%) for categorical variables and mean (SD) for continuous variables unless otherwise stated.

PC, principal component; BMI, body mass index; CVD, cardiovascular disease; HRF, heart rate fragmentation; PIP, percentage of inflection points; IALS, inverse average length of segments; PSS, percentage of short segments; PAS, percentage of alternating segments.

^a^ Comparison across tertiles of principal component 1 using ANOVA Overall F test for continuous variables and χ^2^ test for categorical variables, unless otherwise stated.

^b^ Compared using Kruskal-Wallis test.

**Table S7**. Comparison of Baseline Characteristics between Tertiles of Principal Component 2

|  | | **Low Tertile**  **(< -0.4)**  **(n=200)** | **Middle Tertile**  **(-0.4 to 0.3)**  **(n=201)** | **High Tertile**  **(> 0.3)**  **(n=201)** | ***P-*value^a^** |
| --- | --- | --- | --- | --- | --- |
| PC 2, score | | -1.1 (0.5) | 0.0 (0.2) | 1.1 (0.6) | **<0.001** |
| Age, years | | 73.8 (2.9) | 74.0 (3.0) | 73.6 (2.9) | 0.352 |
| Race, n (%) | |  |  |  | 0.821 |
| White | | 187 (93.5) | 186 (92.5) | 188 (93.5) |  |
| Black | | 10 (5.0) | 12 (6.0) | 8 (4.0) |  |
| Other | | 3 (1.5) | 3 (1.5) | 5 (2.5) |  |
| Education, n (%) | |  |  |  | 0.363 |
| Elementary/Middle School | | 0 (0.0) | 3 (1.5) | 0 (0) |  |
| High School | | 47 (23.5) | 38 (18.9) | 31 (15.4) |  |
| Vocational/Technical School | | 21 (10.5) | 16 (8.0) | 17 (8.5) |  |
| Some College | | 60 (30.0) | 60 (29.9) | 72 (35.8) |  |
| College Degree | | 54 (27.0) | 66 (32.8) | 61 (30.4) |  |
| Post-Graduate College | | 12 (6.0) | 13 (6.5) | 14 (7.0) |  |
| Advanced Degree | | 6 (3.0) | 5 (2.5) | 6 (3.0) |  |
| Smoking Status, n (%) | |  |  |  | 0.997 |
| Current Smoker | | 14 (7.0) | 15 (7.5) | 14 (7.0) |  |
| Former Smoker | | 105 (52.5) | 108 (53.7) | 106 (52.7) |  |
| Never Smoker | | 81 (40.5) | 78 (38.8) | 81 (40.3) |  |
| Drinking Status, days | mean (SD) | 5.9 (9.8) | 6.5 (9.8) | 4.8 (8.7) | 0.184 |
|  | median (IQR) | 0 (5.5) | 0 (10) | 0 (5) | 0.141^b^ |
| BMI, kg/m^2^ | | 28.7 (5.2) | 28.2 (4.8) | 28.6 (4.4) | 0.448 |
| Hypertension, n (%) | | 130 (65.0) | 131 (65.2) | 146 (72.6) | 0.175 |
| Diabetes, n (%) | | 45 (22.5) | 49 (24.4) | 69 (34.3) | **0.016** |
| CVD, n (%) | | 60 (30.0) | 68 (33.8) | 74 (36.8) | 0.350 |
| HRF PIP, % | | 68.3 (8.1) | 68.6 (7.2) | 69.0 (7.0) | 0.617 |
| HRF IALS, beats^-1^ | | 0.7 (0.1) | 0.7 (0.1) | 0.7 (0.1) | 0.610 |
| HRF PSS, % | | 62.6 (9.1) | 62.7 (8.6) | 62.7 (8.2) | 0.998 |
| HRF PAS, % | | 27.4 (9.7) | 27.3 (9.9) | 27.0 (9.9) | 0.919 |

Data are presented as n (%) for categorical variables and mean (SD) for continuous variables unless otherwise stated.

PC, principal component; BMI, body mass index; CVD, cardiovascular disease; HRF, heart rate fragmentation; PIP, percentage of inflection points; IALS, inverse average length of segments; PSS, percentage of short segments; PAS, percentage of alternating segments.

^a^ Comparison across tertiles of principal component 2 using ANOVA Overall F test for continuous variables and χ^2^ test for categorical variables, unless otherwise stated.

^b^ Compared using Kruskal-Wallis test.

**Table S8.** Sensitivity Analysis of the Association between Digital Phenotype of Physical Activity Patterns and Heart Rate Fragmentation Excluding Participants in the Lowest Education Category (n=599)

|  | | **Cluster 2 (Diffuse-Late Phenotype)** | |
| --- | --- | --- | --- |
|  | Model | **β (95% CI)** | ***P-*value** |
| HRF PIP, % | Model 1^a^ | 1.277 (0.074, 2.479) | **0.038** |
|  | Model 2^b^ | 1.251 (0.058, 2.445) | **0.040** |
|  | Model 3^c^ | 1.187 (0.025, 2.349) | **0.045** |
| HRF IALS, beats^-1^ | Model 1^a^ | 0.01277 (0.00077, 0.02478) | **0.037** |
|  | Model 2^b^ | 0.01262 (0.00070, 0.02455) | **0.038** |
|  | Model 3^c^ | 0.01198 (0.00038, 0.02359) | **0.043** |
| HRF PSS, % | Model 1^a^ | 1.263 (-0.131, 2.656) | 0.075 |
|  | Model 2^b^ | 1.061 (-0.298, 2.420) | 0.125 |
|  | Model 3^c^ | 1.025 (-0.314, 2.364) | 0.132 |
| HRF PAS, % | Model 1^a^ | 0.265 (-1.329, 1.858) | 0.743 |
|  | Model 2^b^ | 0.129 (-1.446, 1.703) | 0.872 |
|  | Model 3^c^ | 0.112 (-1.455, 1.679) | 0.888 |

Each heart rate fragmentation outcome was tested separately, with Cluster 1 serving as the reference group.

CI, confidence interval; HRF, heart rate fragmentation; PIP, percentage of inflection points; IALS, inverse average length of segments; PSS, percentage of short segments; PAS, percentage of alternating segments.

^a^ Crude model with twin ID as random intercept.

^b^ Adjusted for age, race, education, BMI, smoking, and drinking, with twin ID as random intercept.

^c^ Adjusted for age, race, education, BMI, smoking, drinking, and history of any composite CVD, with twin ID as random intercept.

**Table S9.** Sensitivity Analysis of the Association between Principal Component Scores and Heart Rate Fragmentation

|  | | **PC 1, score** | | **PC 2, score** | | **Interaction Term** | |
| --- | --- | --- | --- | --- | --- | --- | --- |
|  | Model | **β (95% CI)** | ***P-*value** | **β (95% CI)** | ***P-*value** | **β (95% CI)** | ***P-*value** |
| HRF PIP, % | Model 1^a^ | -0.912 (-1.516, -0.308) | **0.003** | 0.399 (-0.198, 0.996) | 0.189 | 0.212 (-0.366, 0.790) | 0.469 |
|  | Model 2^b^ | -0.508 (-1.145, 0.130) | 0.118 | 0.451 (-0.141, 1.042) | 0.134 | 0.086 (-0.491, 0.663) | 0.768 |
|  | Model 3^c^ | -0.378 (-0.999, 0.244) | 0.232 | 0.313 (-0.265, 0.891) | 0.286 | 0.098 (-0.463, 0.660) | 0.730 |
| HRF IALS, beats^-1^ | Model 1^a^ | -0.00879 (-0.01482, -0.00276) | **0.005** | 0.00397 (-0.00199, 0.00993) | 0.190 | 0.00232 (-0.00345, 0.00809) | 0.429 |
|  | Model 2^b^ | -0.00497 (-0.01134, 0.00140) | 0.125 | 0.00450 (-0.00141, 0.01041) | 0.134 | 0.00113 (-0.00464, 0.00689) | 0.699 |
|  | Model 3^c^ | -0.00362 (-0.00982, 0.00259) | 0.251 | 0.00311 (-0.00266, 0.00888) | 0.289 | 0.00125 (-0.00435, 0.00686) | 0.659 |
| HRF PSS, % | Model 1^a^ | -1.208 (-1.901, -0.515) | **0.001** | 0.136 (-0.559, 0.831) | 0.699 | 0.365 (-0.297, 1.026) | 0.278 |
|  | Model 2^b^ | -0.491 (-1.212, 0.230) | 0.181 | 0.152 (-0.526, 0.830) | 0.658 | 0.179 (-0.474, 0.832) | 0.588 |
|  | Model 3^c^ | -0.390 (-1.103, 0.324) | 0.282 | 0.064 (-0.605, 0.732) | 0.851 | 0.203 (-0.441, 0.848) | 0.534 |
| HRF PAS, % | Model 1^a^ | -0.851 (-1.647, -0.054) | **0.037** | -0.036 (-0.833, 0.762) | 0.930 | -0.085 (-0.845, 0.676) | 0.826 |
|  | Model 2^b^ | -0.291 (-1.126, 0.543) | 0.491 | -0.001 (-0.785, 0.783) | 0.998 | -0.212 (-0.968, 0.544) | 0.580 |
|  | Model 3^c^ | -0.207 (-1.040, 0.626) | 0.624 | -0.062 (-0.844, 0.719) | 0.875 | -0.196 (-0.949, 0.557) | 0.608 |

Each heart rate fragmentation outcome was tested separately, with both principal component scores and interaction term included in models.

PC, principal component; CI, confidence interval; HRF, heart rate fragmentation; PIP, percentage of inflection points; IALS, inverse average length of segments; PSS, percentage of short segments; PAS, percentage of alternating segments.

^a^ Crude model with twin ID as random intercept.

^b^ Adjusted for age, race, education, BMI, smoking, and drinking, with twin ID as random intercept.

^c^ Adjusted for age, race, education, BMI, smoking, drinking, and history of any composite CVD, with twin ID as random intercept.
**Table S10.** Comparison of Model Fit between Phenotype Models and Principal Component Score Models

| Outcome | Phenotype Model AIC | PC Score Model AIC^a^ |
| --- | --- | --- |
| HRF PIP, % | **4036.9** | 4041.1 |
| HRF IALS, beats^-1^ | **-1362.6** | -1339.9 |
| HRF PSS, % | **4195.6** | 4198.5 |
| HRF PAS, % | **4379.2** | 4380.0 |

Each heart rate fragmentation outcome was tested separately.

All models were adjusted for age, race, education, BMI, smoking, drinking, and history of any composite CVD, with twin ID as random intercept.

Smaller AIC values indicate better fit.

PC, principal component; AIC, akaike information criterion; HRF, heart rate fragmentation; PIP, percentage of inflection points; IALS, inverse average length of segments; PSS, percentage of short segments; PAS, percentage of alternating segments.

^a^ Model included principal component 1 score, principal component 2 score, and the interaction term.

**Figure S1.** Horn’s parallel analysis scree plot for principal component retention, with comparisons against the 95^th^ percentiles of resampled and simulated data. PC, principal component.
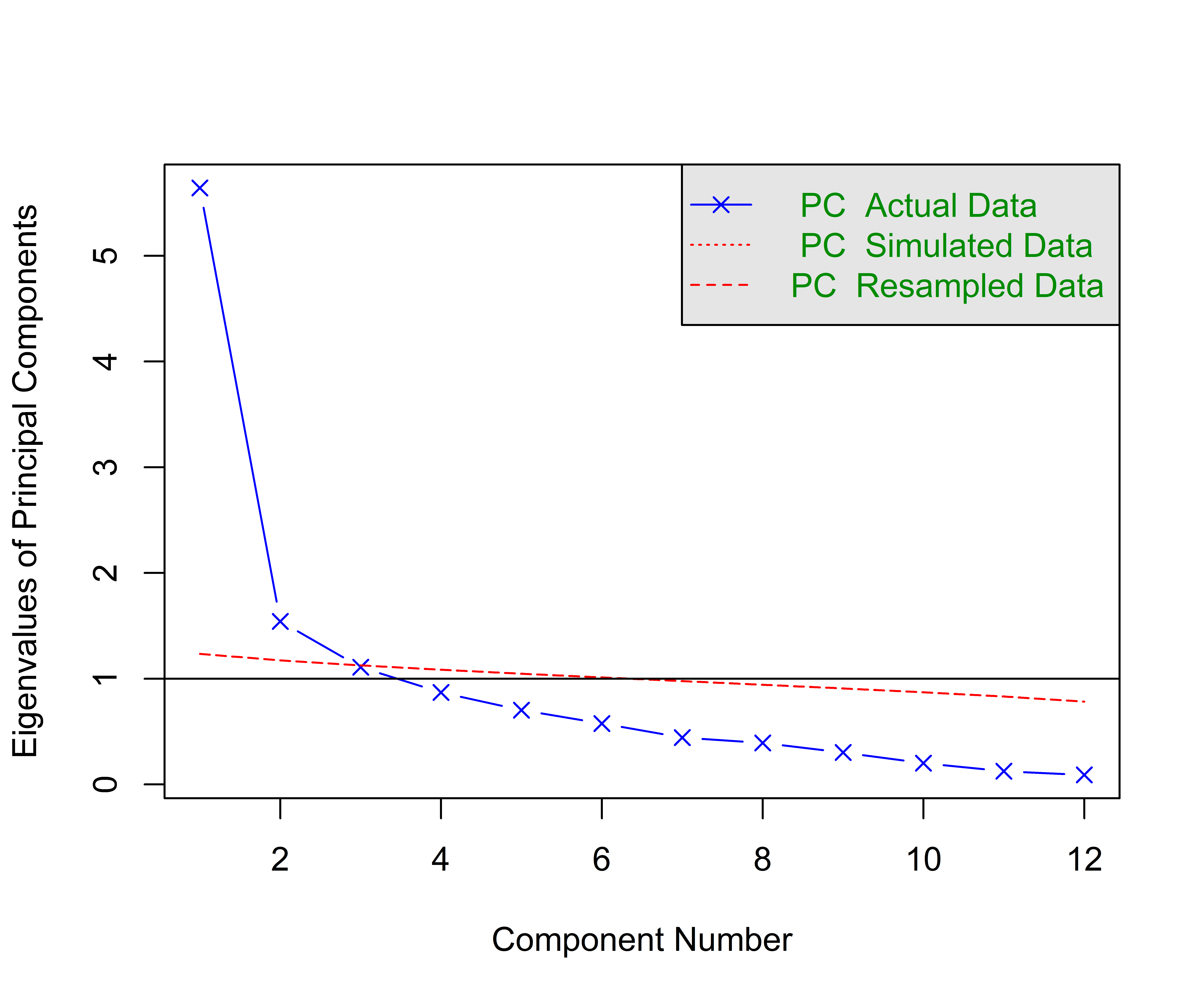

**Figure S2.** Dendrogram derived from agglomerative hierarchical clustering using Ward’s minimum variance method. Height represents the merging cost under Ward’s criterion (the increase in within-cluster sum of squares when clusters are joined). Input data were the two retained principal component scores (n=602).
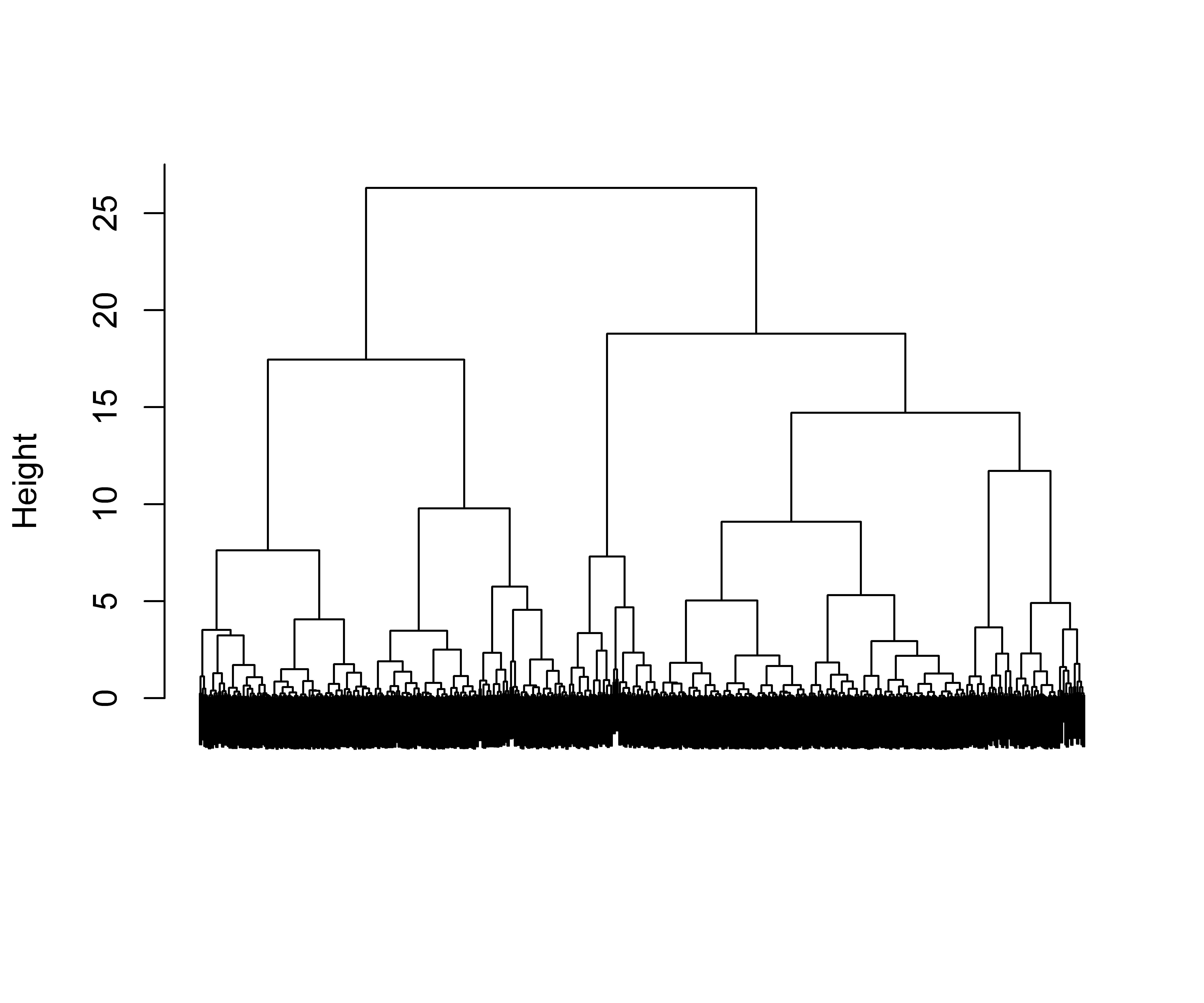
